# Severity of SARS-CoV-2 Omicron XBB subvariants in Singapore

**DOI:** 10.1101/2023.05.04.23289510

**Authors:** Rachael Pung, Xin Peng Kong, Lin Cui, Sae-Rom Chae, Mark I-Cheng Chen, Vernon J. Lee, Zheng Jie Marc Ho

## Abstract

Several XBB subvariants such as XBB.1.5, XBB.1.9, XBB.1.16 and XBB.2.3 co-circulate in Singapore. Despite the different viral properties of XBB.1.16 as compared to other XBB subvariants, comparison on their severity is limited. In this study, we investigate the outcomes of hospitalisation and severe COVID-19 infection in individuals infected with different XBB subvariants, adjusted for potential confounders such as age and vaccination history. Overall, our preliminary analysis showed no difference in the severity of different XBB variants.

## Main

SARS-CoV-2 Omicron sublineage XBB is a recombinant of BA.2.10.1 and BA.2.75 sublineages and was first identified in August 2022 [1]. This strain led to a major outbreak in Singapore which peaked in October 2022 before falling to a low base in early 2023. Since then, several XBB subvariants such as XBB.1.5, XBB.1.9, XBB.1.16 and XBB.2.3 have emerged. While they have similar genetic profiles, XBB.1.16 has been designated as a variant of interest by WHO on 17 Apr, 2023 given its growth advantage and immune escape properties [2, 3]. Unlike previous COVID-19 outbreaks where a sustained rise in cases was attributed to one dominant SARS-CoV-2 variant, the increase in COVID-19 cases in Singapore is now predominantly driven by these four XBB subvariants emerging at different time points but presenting as one confluent outbreak that started in early March 2023. Despite the different viral properties of XBB.1.16 as compared to other XBB subvariants, comparison on their severity is limited. As such, it is unclear whether increases in the number of hospitalised cases is an outcome of faster transmission or higher severity or both. In this study, we investigate the outcomes of hospitalisation and severe COVID-19 infection in individuals infected with different XBB subvariants, adjusted for potential confounders such as age and vaccination history.

In Singapore, whole genome sequencing (WGS) is performed on a subset of confirmed COVID-19 cases (both locally infected and imported cases) who tested positive via PCR test administered by a healthcare provider. From 1 January to 18 April 2023, WGS was performed on respiratory samples from 5,518 cases of which 3,798 were infected with an XBB subvariant (Table 1) — 527 (14%) were XBB.1.16, 965 (26%) were XBB.1.5, 920 (24%) were XBB.1.9, 698 (18%) were XBB.2.3, and the remaining 688 (18%) were other XBB sublineages (Table 1).

**Table 1.**
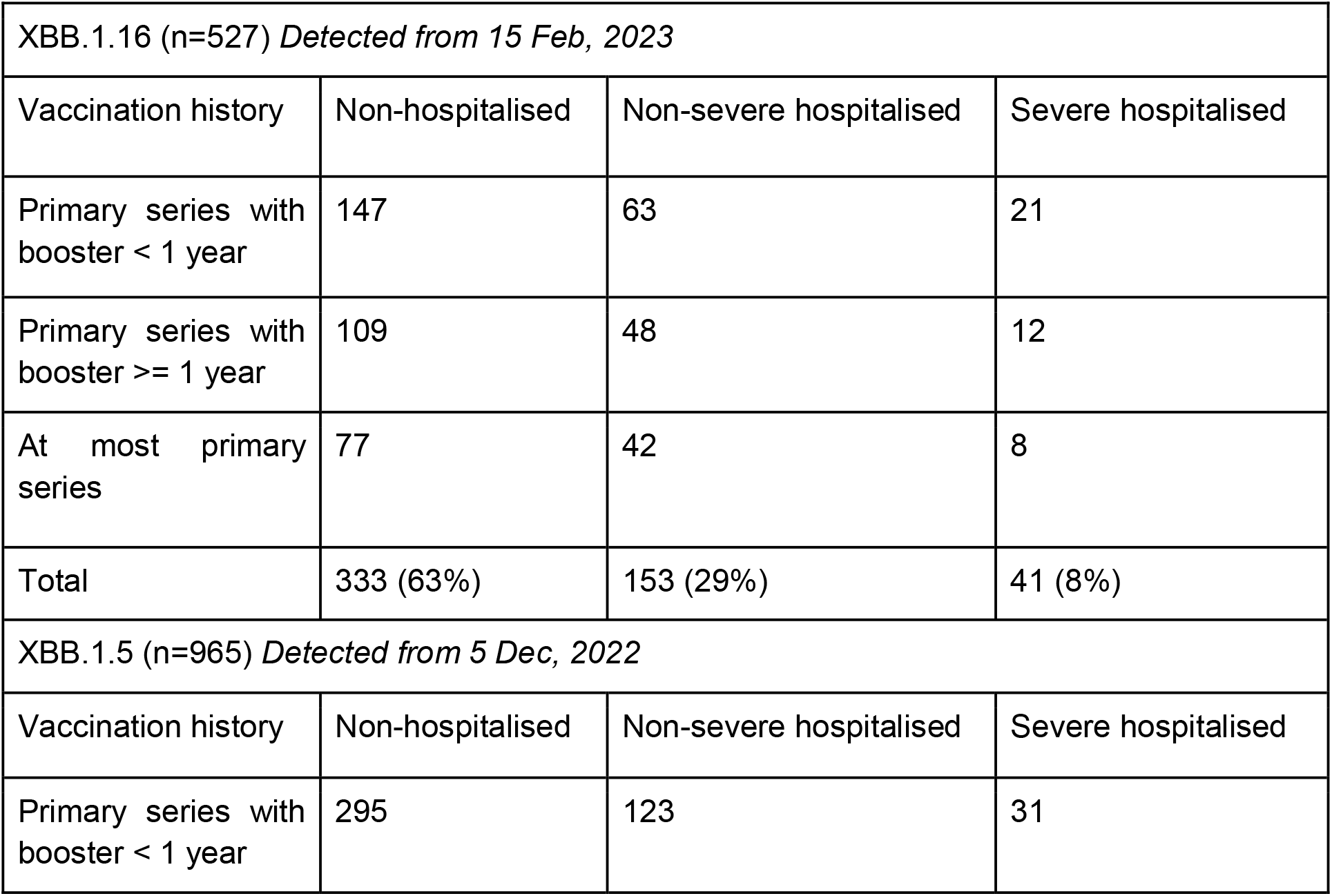
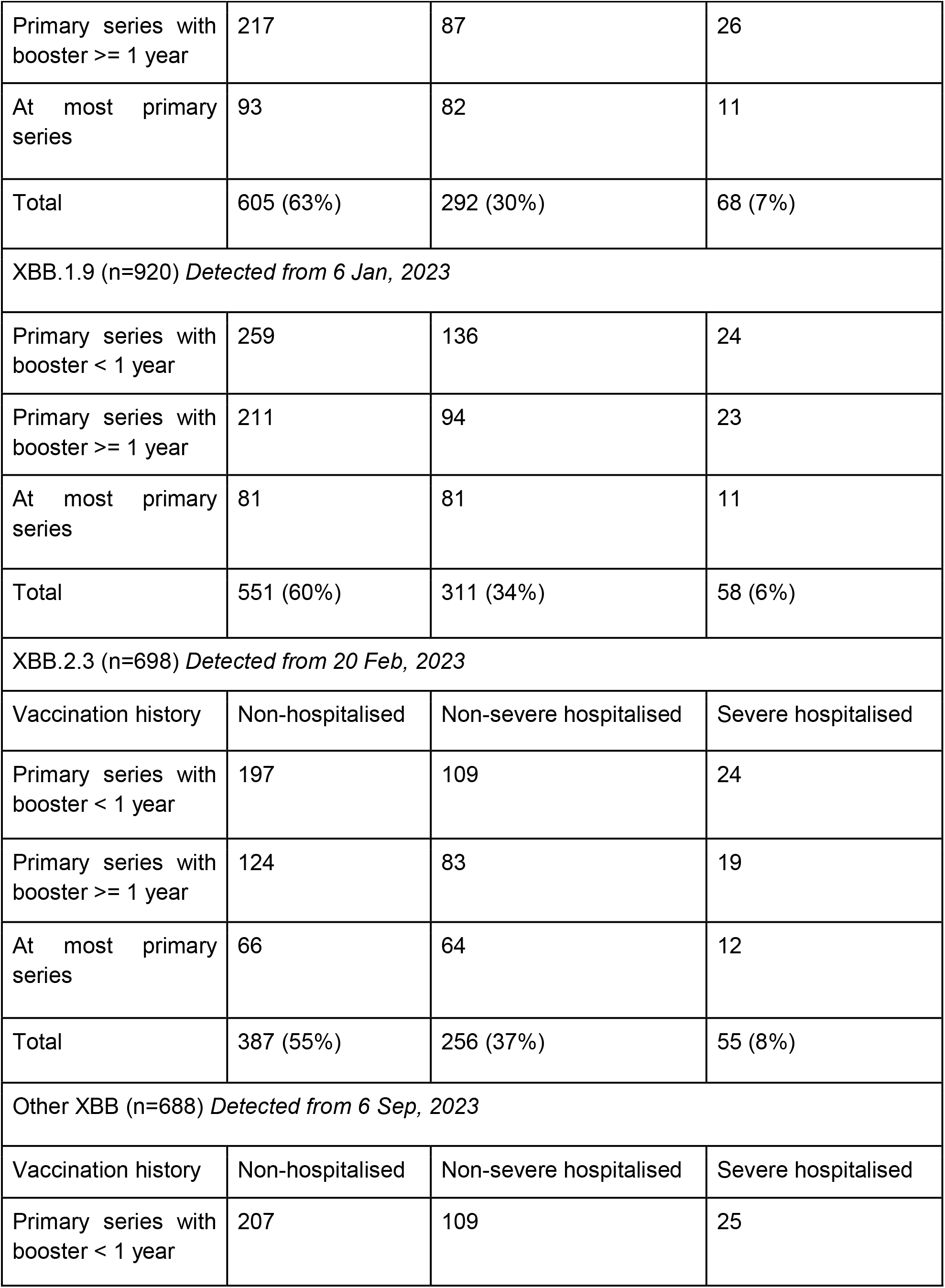
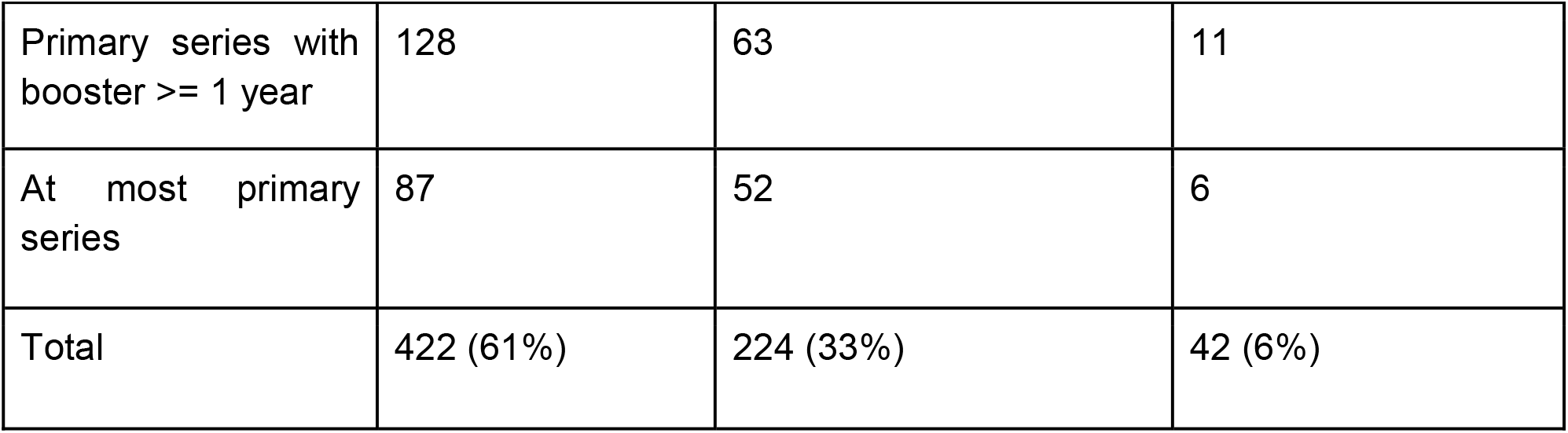
Characteristics of XBB cases by vaccination history and clinical outcomes

Multivariate logistic-regression was used to investigate the (i) severity and (ii) outcomes of hospitalisation due to infection with different SARS-CoV-2 Omicron XBB subvariants, after adjusting for age and vaccination history. A severe infection is defined as a hospitalised case who either required supplemental oxygen, was admitted to ICU or died, while a non-severe infection is defined as all other hospitalised or non-hospitalised cases. The vaccination history of a case is classified as (i) completed the primary vaccination series and had a booster in less than 1 year prior to the current infection episode, (ii) completed the primary vaccination series and had a booster 1 year or more prior to the current infection episode, (iii) at most completed the primary vaccination series.

After adjusting for the confounders, we observed no significant differences in the severity of COVID-19 infection or hospitalisation outcomes across different XBB subvariants (Figure 1). Regardless of the XBB subvariant of infection, the risk of hospitalisation was about 1.5 times higher in those who at most completed their primary vaccination as compared to those with primary vaccination and a booster (Figure 1B and 1D). Given that vaccination history of imported cases may be incomplete, sensitivity analysis was performed using local cases only and similar findings were observed.

**Figure 1.**
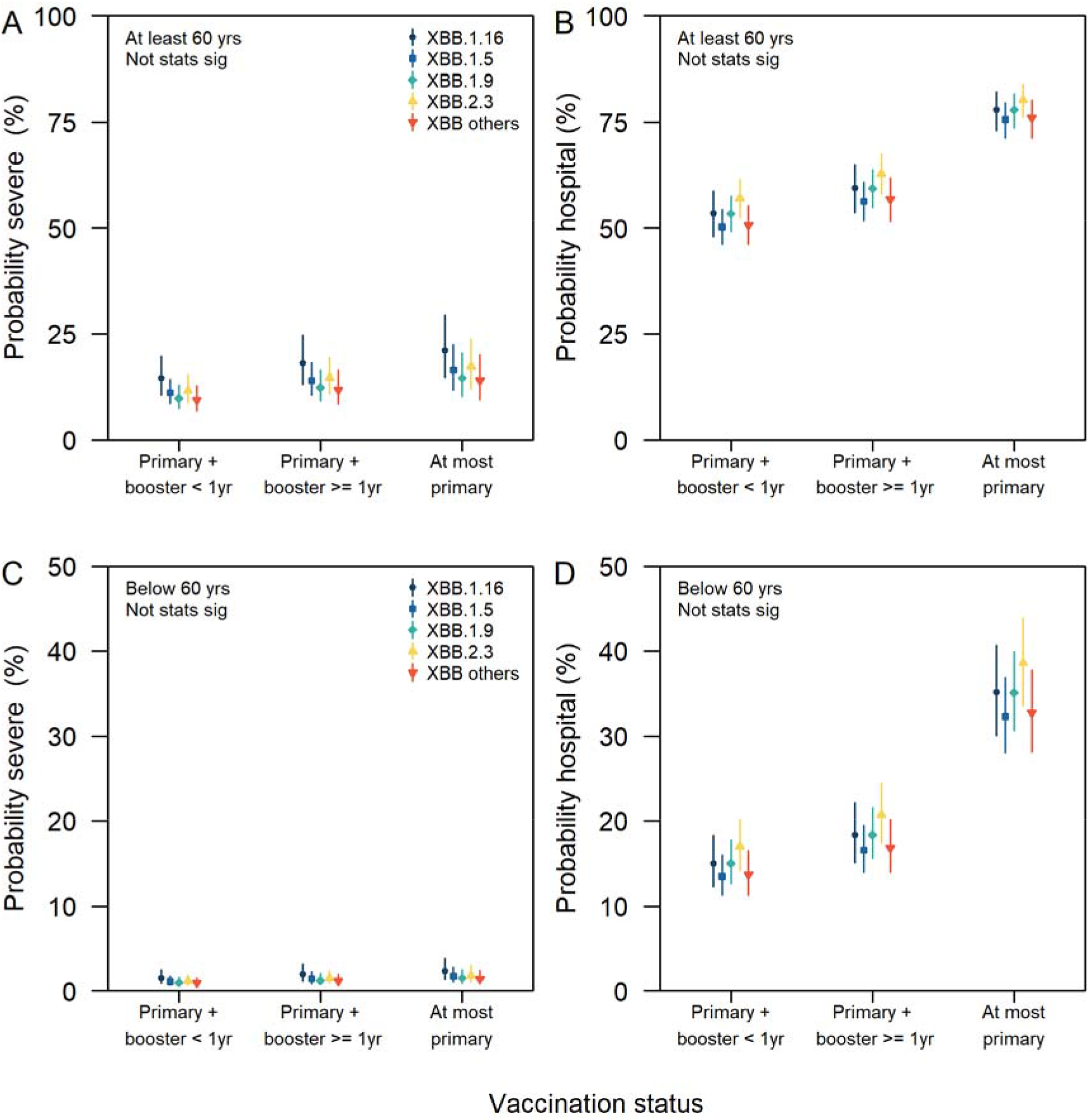
Probability of infection outcomes for different XBB subvariants and vaccination history; (A) severe infection in those aged 60 and above, (B) hospitalisation in those aged 60 and above, (C) severe infection in those aged below 60, (D) hospitalisation in those age below 60. Results were not statistically significant.

Our analysis has some limitations. Given the low number of hospitalised COVID-19 cases, a higher proportion (60-90%) of these cases were sampled for WGS. In contrast, about 10% of the non-hospitalised cases were sampled for WGS. However, this sampling criteria is consistent across the period of study. Accounting for this sampling bias would increase the proportion of non-hospitalised cases in Table 1, but would not affect the mean outcomes of the multivariate logistic regression. Delays in the onset of severe infection may bias the severity of XBB.1.16 and XBB.2.3 downwards, given that these two variants were only detected in samples from mid Feb 2023 onwards. In the current dataset, about 50% of the hospitalised XBB.1.16 and XBB.2.3 cases were notified before Apr, while this was 60% in the non-hospitalised XBB.1.16 and XBB.2.3 cases. Thus, the impact of delayed outcome is not likely to have had a large effect on the analysis.

Overall, our preliminary analysis showed no difference in the severity of different XBB variants. As countries scale down on COVID-19 testing and reporting, studies to distinguish the growth advantage of different COVID-19 variants would be increasingly challenging given the drop in case ascertainment, lack of contact exposure data and studies on the impact of hybrid immunity on the risk of transmission [4]. However, continued data collection on the clinical severity of cases and such comparative analyses would help to identify changes in severity of SARS-CoV-2 variants and hence factors contributing to the growth in COVID-19 hospitalisations.

## Data Availability

All data produced in the present work are contained in the manuscript

## Ethics

The study was approved by the London School of Hygiene & Tropical Medicine Observational Research Ethics Committee (ref. 25727). All data and analysis were collected and performed in line with the Infectious Diseases Act in Singapore which permits the collection and publication of surveillance data.

